# External Causes of Death in Sulaymaniyah City in 2022

**DOI:** 10.1101/2025.01.25.25321120

**Authors:** Kawan Rasul, Hemn Hassan, Eram Abdalla, Farman Tahir, Sozyar Ahmed, Xalat Reza, Payam Faraj, Lana Hamaamin

## Abstract

**Background:** External causes of death like injuries, violence, and accidents have a great impact on population and depletion of health resources. Accurate and precise data on external causes is important for informing public health policies. This study analyzed the most common external causes of death in Sulaymaniyah city during 2022 and examined characteristics of the deceased individuals.

**Methods:** This cross-sectional study included 499 cases of death from external causes in Sulaymaniyah in the year 2022. These data were obtained from the Medico-Legal Institute. Descriptive statistics, cross-tabulations, and chi-square tests were performed to find out common causes, sex differences, and age patterns.

**Results:** Out of 499 cases, road traffic accidents (36.1%) were the most common cause, followed by burn injuries (17.2%) and gunshot wounds (16.8%). Males predominated with a 2.4:1 ratio over females. Road accidents were more common among males, while burn injuries were higher in females. All age-groups were affected mainly by road traffic accidents, and the age group of 25-44 years was most affected. Burn injuries and gunshot wounds were prevalent causes across age-groups 15-44.

**Conclusion:** Road traffic accidents, burn injuries, and gunshot wounds were the major external causes of death in Sulaymaniyah in 2022, with distinct sex and age patterns. Findings suggest prompt needs for improving road safety, applying burn prevention strategies, and planning interventions for vulnerable groups to reduce this preventable mortality burden

## 1. Introduction

### 1.1 Background

Injuries, violence, and accidents are leading external causes of death (ECsD) highly related to mortality globally. These ECsD have major social and economic impacts. According to the World Health Organization estimates, ECsD have a huge impact on the health of populations, causing human suffering and depleting health system resources (WHO, 2014). Moreover, road traffic accidents, falls, and violence are responsible for 9% of all deaths (WHO, 2016).

Low and middle-income countries have much higher mortality rates due to external causes, for example, some of the Middle East countries. For example, in Iraq, it was reported that some of the highest rates of these incidents are in this region, as reported by Smith et al. (2020). However, most of the research done in Iraq is focused on the south and central region, leaving the Kurdish region underrepresented. Some anecdotal evidence and medical reports show the increased burden of ECsD in Sulaymaniyah, but even these two prominent forms, namely RTAs and burns, have not been properly statistically recorded as yet. Without solid data to work with, we’re essentially working in the dark when trying to tackle this critical issue. This gap in our understanding makes it clear - we need to focus our research efforts on understanding these external causes of death (ECsD) in our community.

### 1.2 Problem definition

Sulaymaniyah is a city in the Kurdistan Region of Iraq. In the year 2022, it showed a remarkable number of deaths due to external causes. The five most important external causes of death among people living in Sulaymaniyah include burn-related deaths, drowning, electrocution, firearm incidents, and traffic accidents. External causes of death imposed a substantial burden in the Kurdistan region and Sulaymaniyah in particular, since it is stated that more than 9% of mortality globally is caused by external factors (WHO, 2016). Based on that, precise and accurate data about the specific external causes, apart from demographic information for those fatalities, is a crucial and initial step to inform and regulate public health policies. Apart from these, fatal injuries result in very high economic costs regarding medical expenses, lost wages/productivity, and decreased workforce participation. According to the WHO estimates, road traffic crashes alone have been estimated to cost approximately 3% of most countries’ GDP (WHO, 2004). The understanding of specific causes, demographic influences, and associated risk factors facilitates evidence-based formulation and implementation of policies and interventions by the public health agencies with particular attention to high-risk groups. Accurate surveillance data are needed to clearly delineate the hotspots and vulnerable populations that need targeting for prevention. (Chandran et al., 2010).

### 1.3 The Importance of the Study

this study is important, as it will provide essential information about the epidemiology of ECsD in the Sulaymaniyah city. This will be able to provide evidence for public health policy and interventions aimed at reducing premature mortality and economic and social burdens associated with ECsD by focusing on the most common ECsD, their gender-specific patterns, and age-group variations. By focusing on Sulaymaniyah city, this research will help in identifying high-risk groups and assist in developing preventative measures accordingly.

### 1.4 Research Questions

1. What are the main external causes of death in Sulaymaniyah city in the year 2022?
2. Are there any statistically significant differences in the external causes of death by gender in Sulaymaniyah?
3. How do the external causes of death vary across different age groups in Sulaymaniyah?
4. What are the demographic patterns, including gender and age, of the leading causes, such as road traffic accidents, burn injuries, and gunshot wounds?
5. Are there any age groups or genders with higher susceptibility to certain external causes of death in Sulaymaniyah?

### 1.5 Aim and Objectives of the Study

The present study discussed and explored the most predominant external causes of deaths in Sulaymaniyah city during 2022, further deliberation upon the nature of the victims. Key objectives of this research are:

1. To identify the most common ECsD
2. To study gender-specific trends in the presentation of ECsD
3. To identify the age-group variations concerning common ECsD

## 2. Methodology

### 2.1 Study design

a cross sectional study design is chosen because it is suitable for determining prevalence and patterns of external causes of death.

### 2.2 Study setting and duration

the study was conducted in Sulaymaniyah city, Kurdistan region of Iraq. Which is a large city and has a population about 2 million people. There were 698 cases of people who have died due to external causes from 1^st^ of January 2022 to 31^st^ of December 2022 in Sulaymaniyah city which the data was collected in Medico-Legal institute which is the main faculty responsible for maintaining official data of death caused externally.

### 2.3 Inclusion Criteria

All recorded deaths from external causes in 2022 (n=698). Among them cases with complete information such as (cause of death, age, gender and date of birth) were included.

### 2.4 Exclusion Criteria

out of 698 initial cases some of them were excluded because some internal causes of death misclassified as external like Myocardial Infarction, some missed data, and some had incomplete demographic information. The final sample size was 499 cases (71.5% of initial cases).

### 2.5 Data collection

As we used the documented and available data from Sulaymaniyah Medico-Legal Institute. So, the electronic and finalized data were obtained through a copy of the excel dataset. Additionally, variables collected including age, gender, primary cause of death, date of death, and place of death. We cleansed the dataset for incorrect, irrelevant, incomplete and missed data to make sure it is accurate and suitable for analysis.

### 2.6 Classifications

some variables needed classifications for example age and it was classified into Children (0-14 years), Young adults (15-24 years), Adults (25-44 years), Middle-aged (45-64 years) and Elderly (≥65 years). Adding to that, causes of deaths were classified into Car accidents, Burn injuries, Gunshot wound, Drowning, Motorcycle accidents, fall injuries, Electrocution and Other causes (blast injuries, blunt force trauma, etc.)

### 2.7 Study tool

Because of lack of direct and/or indirect contacts with patient/client(s) as we work with about dead people and specifically with their recorded data, so we didn’t need any tool/objective(s) to conduct our research.

### 2.8 Statistical Analysis

We obtained an excel dataset from Sulaymaniyah Medico-Legal institute, we checked the dataset for errors or missing and non-applicable data and we corrected them. Afterward, we transformed the Excel Document into [Statistical Package for Social Science-IBM SPSS/version 25] for data analysis, whereupon we obtained all descriptive statistics like frequencies and percentages for each variable and presented them as figures, charts and tables. Furthermore, inferential statistics have been found by employing particular tests to describe the correlations and associations between variables, such as (Cross Tabulation and Chi-Square) and significance level set at p. value <0.05.

### 2.9 Ethical Considerations

Prior to initiation, we first obtained official permission out from the dean of the College of Health and Medical Technology/Nursing Department/ Sulaymaniyah Polytechnic University (SPU), followed by approval from the Sulaymaniyah Directorate of Health. Also, before the official beginning of the experiment, authorization was acquired from other appropriate authorities include Director of Sulaymaniyah Medico-Legal Institute, Sulaymaniyah Statistics Directorate and the Registration Bureau of Births & Deaths.

The collected data does not have names, addresses, contact information, or any specific information that allow us to track the deceased, so the data was completely anonymous. This level of anonymity ensures complete privacy protection of the deceased and their families

## 3. Results

### 3.1 Descriptive statistics

All the collected data on the ECsD during 2022 was 698 cases. After exclusion of the non-applicable data for this study, the remaining was data of 499 cases. This dataset consists of several demographic variables and ECsD.

The dataset shows that Sex distribution is asymmetrical; since male cases were predominant with 353 and 146 females, i.e. male to female ratio with 2.4:1 as showed in Figure 1.

**Figure 1:**
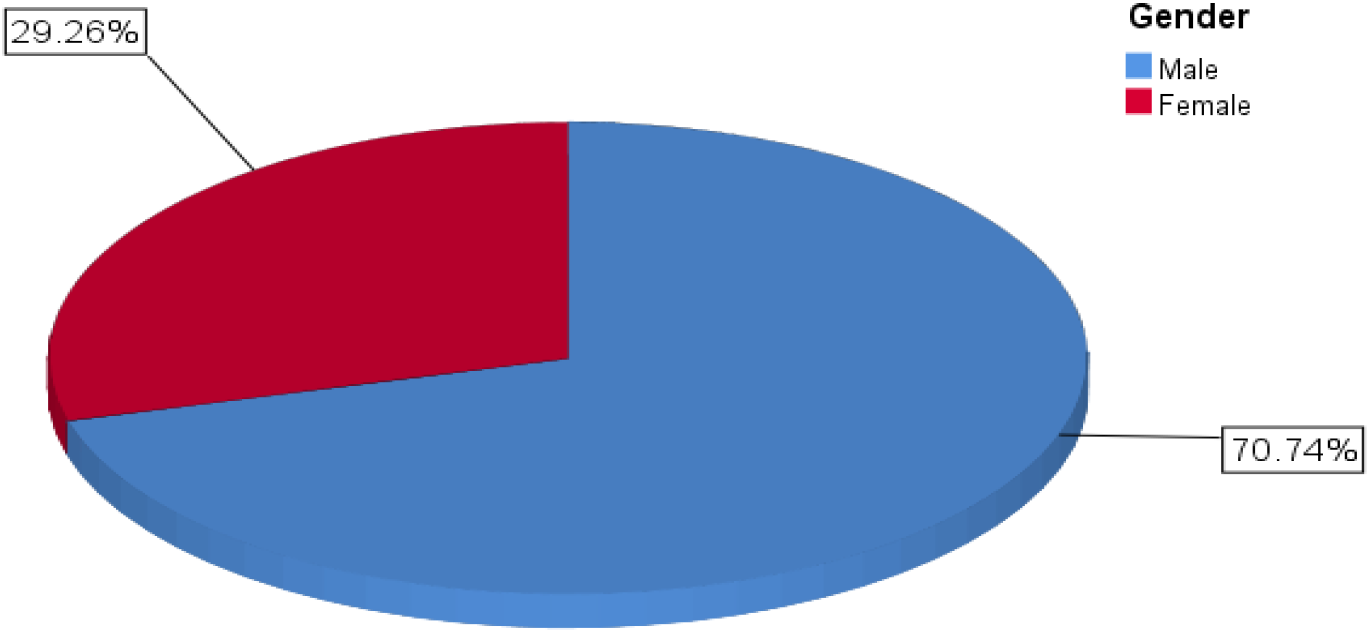
Male to female ratio.

Additionally, traumatic causes of death accounted for 12.5% of total deaths and non-traumatic causes encompassed 87.5% of total deaths in Sulaymaniyah city for 2022.

Following that, the dataset demonstrates that people who are at age (15-44) are more frequently affected specifically ages of (25-44). As it demonstrated in Figure 2.

**Figure 2:**
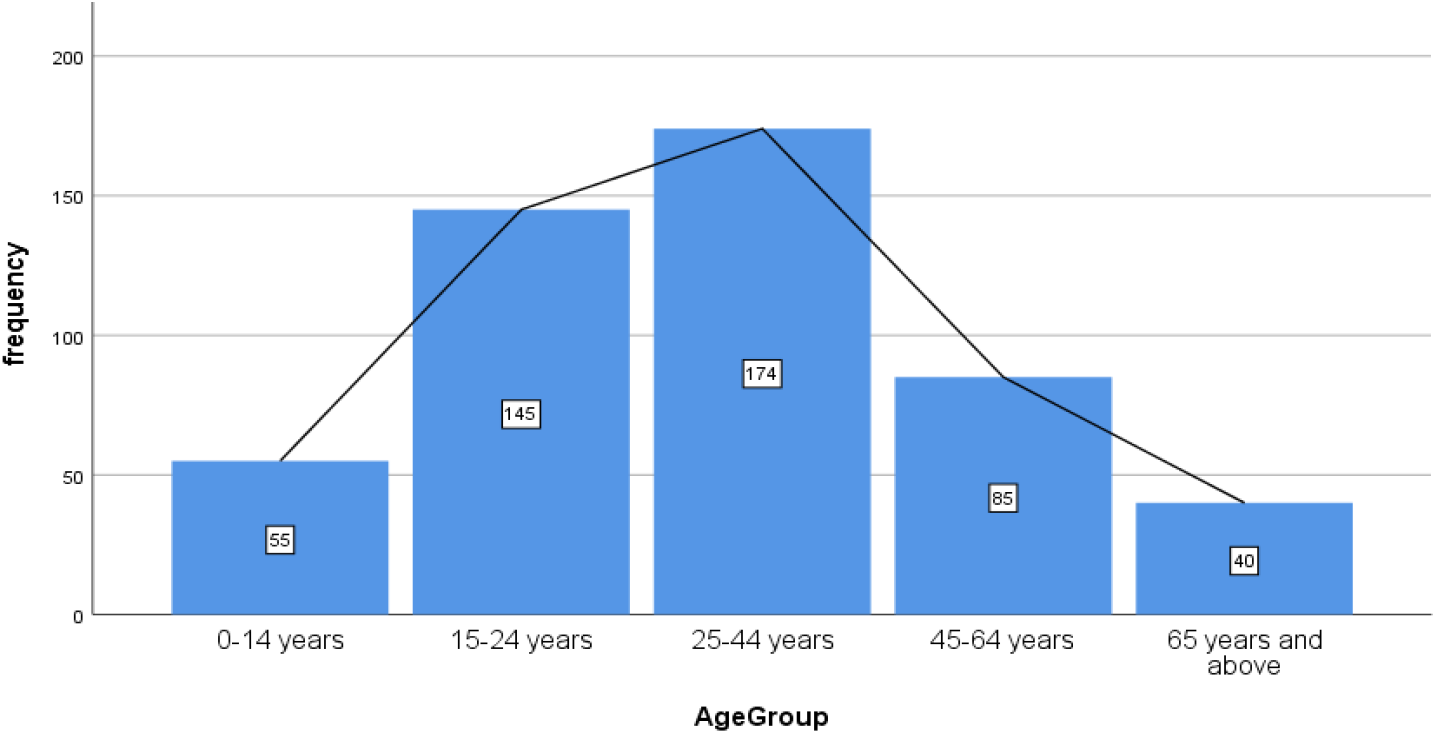
Age-group distribution.

Furthermore, as it is shown in Figure 3, the number of deaths most frequently registered due to external causes were in the months of May, June and July primarily July. To be noted, in the remaining months external deaths are relatively similar.

**Figure 3:**
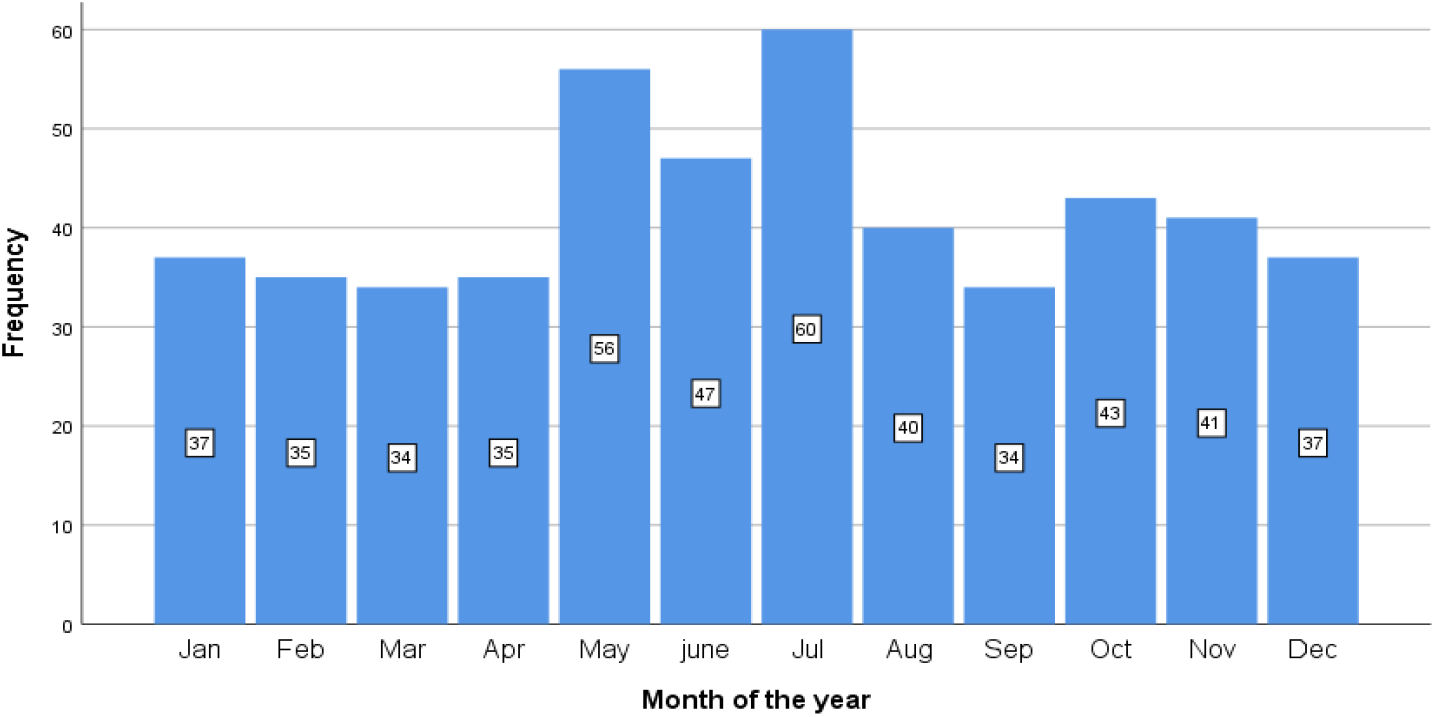
Distribution of death registration per month.

### 3.2 Most common (ECsD)

RTA, which encompasses both car accidents and motorcycle accidents accounted for 36.1% of all causes of death with the frequency of 180, followed by burn injuries and gunshot. Additionally causes like fall injuries, electrocution and others were less frequently occurred (less than 10%), as shown in the Table 1.

**Table 1:** Common ECsD.

|  |  | Frequency | Percent |
| --- | --- | --- | --- |
| Valid | Car accidents | 145 | 29.1% |
|  | Burn injuries | 86 | 17.2% |
|  | Gunshot wound | 84 | 16.8% |
|  | Drowning | 51 | 10.2% |
|  | Motorcycle accidents | 35 | 7.0% |
|  | Fall injuries | 34 | 6.8% |
|  | Electrocution | 30 | 6.0% |
|  | Other causes (blast injuries, blunt force trauma, etc.) | 34 | 6.8% |
|  | Total | 499 | 100.0 |

### 3.3 Sex-specific common ECsD

Sex played an important role in shaping which cause of death is more frequent in each Sex. As the following table shows, there are different patterns of Sex and causes of death. For instance, RTA account for 151 cases which equals to 42.7% of all external causes in males. However, in female study population RTA is less predominant and accounted for 17.8% of all external causes. The detail is shown in the Table 2.

**Table 2:** Sex-specific common causes of death.

|  |  | Sex |  |  |  |
| --- | --- | --- | --- | --- | --- |
|  |  | Male | Female |  | Total |
| Cause of death | Car accidents | 119 | 26 |  | 145 |
|  | Burn injuries | 21 | 65 |  | 86 |
|  | Gunshot wound | 69 | 15 |  | 84 |
|  | Drowning | 37 | 14 |  | 51 |
|  | Motorcycle accidents | 32 | 3 |  | 35 |
|  | Fall injuries | 24 | 10 |  | 34 |
|  | Electrocution | 23 | 7 |  | 30 |
|  | Other causes (blast injuries, blunt force trauma, etc.) | 28 | 6 |  | 34 |
|  | Total | 353 | 146 |  | 499 |

Its significance is tested using Pearson chi square test which it ensures its significance with p-value equals to 1.74*10^−19^ which is greatly less than the significant p-value (0.05).

### 3.4 Age-group patterns of common ECsD

Some age-groups were more vulnerable and some age-groups possess similar causes of deaths. RTA (36.4%) and burn related deaths (22%) were more profound in the first age-group (0-14 years). Moving to the second age group (15-24 years), RTA accounted for (35%) of all external deaths followed by burn injury related (21.4%), gunshot related (18%) and drowning (14%). The details of age-groups are demonstrated in the Table 3. The significance of findings is tested using Pearson chi-square and it proved its significance with a p-value equals to 0.001.

**Table 3:** Age-group variations of external causes.

|  |  | Age-Groups |  |  |  |  | Total |
| --- | --- | --- | --- | --- | --- | --- | --- |
|  |  | 0-14 years | 15-24 years | 25-44 years | 45-64 years | 65 years and above |  |
| Cause of death | Car accidents | 16 | 32 | 41 | 38 | 18 | 145 |
|  | Burn injuries | 12 | 31 | 29 | 8 | 6 | 86 |
|  | Gunshot wound | 4 | 26 | 35 | 15 | 4 | 84 |
|  | Drowning | 5 | 20 | 20 | 5 | 1 | 51 |
|  | Motorcycle accidents | 4 | 19 | 9 | 1 | 2 | 35 |
|  | Fall injuries | 4 | 6 | 12 | 5 | 7 | 34 |
|  | Electrocution | 2 | 5 | 16 | 5 | 2 | 30 |
|  | Other causes (blast injuries, blunt force trauma, etc.) | 8 | 6 | 12 | 8 | 0 | 34 |
|  | Total | 55 | 145 | 174 | 85 | 40 | 499 |

## 4. Discussion

### 4.1 Overview of External Causes of Death

Traumatic causes accounted for 12.5 percent of total deaths in Sulaymaniyah in 2022 according to the data collected from the Medico-Legal institute in Sulaymaniyah. This was higher than the 9 percent reported in the United States for the year 2017 (Peterson, 2020). Thus, this suggests that the occurrence of traumatic conditions was higher in Sulaymaniyah. Differences in health services, infrastructure, and measures such as road safety and political and social environments may be responsible for such variations.

### 4.2 Gender Patterns in External Mortality

The result of this research revealed that the rate of death from external causes is higher in males than in females at a rate of 71% and the difference was statistically significant (p value<0.001). Additionally, this was supported by a report from Al-Hadithi et al., (2019) which states that males are more at risk from experiencing deaths caused externally. The logic behind this might be attributed to cultural factors in Kurdish society as far as gender concerns are involved, as well as the fact that a major number of males involve in risky activities.

### 4.3 Road Traffic Accidents: Leading Cause of External Mortality

Furthermore, it is revealed that out of all external causes of mortality, RTAs ranked first as the primary mortality cause, contributing to 36.1% of external deaths, and males experienced a higher percentage, 77.5%. This is also a comparable finding to that of Shaho et al. (2020), in which RTAs accounted for 37% and 36% of external deaths in 2012 and 2013, respectively, within Sulaymaniyah. This may also be attributed to the fact that the majority of drivers in Sulaymaniyah city are male and they tend to indulge in risky driving behaviors such as over speeding and breaking traffic rules (Rhodes & Pivik, 2011).

### 4.4 Burn-Related Mortality: A Gender-Specific Challenge

Burns were the most common cause of death in females, even more common than RTAs, out of 76% of external deaths in females was burn-related. Raheem (2012) supports that finding who stated that 61% of burn injuries out of 884 cases in Sulaymaniyah plastic and burn hospital were females. This heightened risk of burn-related injuries in females was linked to some regional factors such as women are more exposed to cooking related hazards, since they are the expected person to cook and prepare meals. Additionally, self-immolation is higher within females in several Asian countries and Middle East.

### 4.5 Age-Related Patterns

The outcome of the research provided insights on ECsD relation with age. Based on the collected data, we found out that some age groups are more at risk for external causes of death. For example, the age-group which is the most frequently affected was 15-44 and more specifically 25-44. Also, WHO, (2020), shows similar patterns in Iraq, Iran and Turkey. This increased mortality can be related to the likelihood from this group to partake in high-risk activities like reckless driving and violence which in turn add a significant risk for external deaths (Rhodes & Pivik, 2011b).

### 4.6 Age-Specific Vulnerabilities

The findings concluded that, RTA was the most common cause of death in all the age-groups. Across the globe, RTAs result in serious health concerns and significant economic burden, it is estimated that more than 1.35 million people are killed or disabled by traffic accidents (Ahmed et al., 2023), according to the same source, in 2019, 93% road traffic related mortality happened in low and middle income countries with an estimated burden of 1.3 million deaths.

Moreover, children aged 0-14 was particularly affected by RTA (36.4% of all external deaths of children), and burn-related was 22%, this may be due to age-related factors like developmental limitations and physiological vulnerability to injuries (Hyder et al., 2008). The pattern remained the same for 15-65 age groups as RTA consistently was the predominant cause followed by Burn-related deaths, except a small distinction in 25-44 age-group which gunshot-related mortality 20% was slightly more common than Burn-related 17%. Lastly, as for the elderly population (>65years), RTA accounted for half of external deaths followed by fall-injuries (17.5%), this finding aligns with regional mortality data in Erbil, where it was demonstrated that injuries and accidents are constituted to 29% of deaths in this age group including internal causes (Emhj, 2017). The high proportion of RTA in this age group requires particular attention and may be related to age-specific vulnerabilities that need specialized interventions.

### 4.7 Seasonal Variations

Our finding suggested a seasonal pattern regarding external causes of death, which peak external mortality was in May and July particularly July. This pattern may be related to increase of outdoor activities and tourism-related traffics. However, more research is necessary to understand the factors that are responsible for this seasonal variations.

### 4.8 Study Limitations

1. The study was done using cross-sectional method, which prevents us from finding causal relationships.
2. The exclusion of around 30% percent of data which might affect the representativeness of our sample.
3. We collected only a single years’ worth of data, so it may not be so effective in capturing long term trends and unusual events.
4. Potential misclassification of causes of death by the institute, since we witnessed some major mistakes like classifying heart attack as an external cause of death.

### 4.9 Future Research Directions

1. Longitudinal studies to track trends over multiple years.
2. Detailed analysis on seasonal patterns in External Mortality.
3. The classification methods and rules used by Medico-Legal institute in relation to External Causes of Death.
4. Assessment of the correctness of data entry and correct classification of external causes of death by local authorities
5. Socioeconomic determinants and their correlation with External mortality like occupation and monthly income

## 5. Conclusion

In This study examined the most common ECsD in Sulaymaniyah city during 2022 with the analysis of patterns by sex and age group. Several key findings came from the analysis of 499 cases. It was found that, RTA being the leading cause followed by burn injuries and gunshot wounds were the next most common ECsD. Notable Sex differences were seen, with RTA accounting for the highest number of external deaths among males but not females. Contrary, burn injuries were the top external cause for females. Gunshot wounds were more common among males than females. Age patterns were also observed. RTA and burn injuries were the top two causes across most age-groups, except among those 65 and older where falls surpassed burn-related deaths. Gunshot wounds was the third common cause for ages 15-64. Drowning were most common in the 15-24 and 25-44 age ranges. The findings emphasize on implementing plans to increase road traffic safety, fire prevention, and limiting firearm availability for people unless licensed and strengthen the requirements. They are key areas for intervention to reduce premature deaths from external causes in Sulaymaniyah. Adding to that, efforts based on the age and Sex patterns can enhance the effectiveness of these prevention programs.

## 6. Recommendations

Based on our findings there are several points we suggest in order to reduce external causes mortality

1. Implementing strategies which ensure road safety which includes fixing roads, setting speed camera limits and disseminate awareness for people especially drivers.
2. Initiating a comprehensive plan to reduce burn-related injuries these include Public education and awareness and Legislations for product safety improvements
3. Planning on restricting firearm availability and accessibility by strengthening conditions to get licensed and preventing illegal weapon stores.

## Data Availability

All data produced in the present study, including the used dataset, is available upon request.

## References

Ahmed, S. K., Mona Gamal Mohamed, Salar Omar Abdulqadirb, Rabab G. Abd El-Kader, El-Shall, N. A., Chandran, D., Ebad, M., & Kuldeep Dhama. (2023). Road traffic accidental injuries and deaths: A neglected global health issue. Health Science Reports, 6(5). 10.1002/hsr2.1240

Ari Raheem, M. (2012) ‘Burn admissions in a teaching hospital in Sulaymaniyah, Iraq’, Annals of Burns and Fire Disasters, 25(3), p.131. Available at: https://www.ncbi.nlm.nih.gov/pmc/articles/PMC3673832/

Cleary, M., Singh, J., West, S., Rahkar Farshi, M., Lopez, V., & Kornhaber, R. (2021). Drivers and consequences of self-immolation in parts of Iran, Iraq and Uzbekistan: A systematic review of qualitative evidence. Burns, 47(1), 25–34. 10.1016/j.burns.2019.08.007

Hyder, A. A., Muzaffar, S. S. F., & Bachani, A. M. (2008). Road traffic injuries in urban Africa and Asia: A policy gap in child and adolescent health. Public Health, 122(10), 1104–1110. 10.1016/j.puhe.2007.12.014

Mifsud, D., Attard, M., & Ison, S. (2017). Old Age: What are the Main Difficulties and Vulnerabilities in the Transport Environment? Transport and Sustainability. 10.1108/s2044-994120170000010017

Al-Hadithi, T., Sharma, S. and Almustafa, M. (2019) ‘Mortality from External Causes in Iraq’, Journal of Forensic Sciences & Criminal Investigation, 12(5). 10.19080/JFSCI.2019.12.555849

Shaho, C., Mohammed, O., Abdulkarim, A.Z. et al. (2020) ‘RTA in Sulaymaniyah province of Iraq’, International Journal of Community Medicine and Public Health, 7(7), pp.2752–2756. 10.18203/2394-6040.ijcmph20202657

Peterson, A. B. (2020). Deaths from Fall-Related Traumatic Brain Injury — United States, 2008–2017. MMWR. Morbidity and Mortality Weekly Report, 69(9). 10.15585/mmwr.mm6909a2

Rhodes, N., & Pivik, K. (2011a). Age and gender differences in risky driving: The roles of positive affect and risk perception. Accident Analysis & Prevention, 43(3), 923–931. 10.1016/j.aap.2010.11.015

emhj (2017) Mortality trends in Erbil, Iraq, 2007–2011. Available at: https://www.emro.who.int/emhj-volume-25-2019/volume-25-issue-5/mortality-trends-in-erbil-iraq-20072011.html

Rhodes, N., & Pivik, K. (2011b). Age and gender differences in risky driving: The roles of positive affect and risk perception. Accident Analysis & Prevention, 43(3), 923–931. 10.1016/j.aap.2010.11.015

World Health Organization (WHO) (2013) Global Status Report on Road Safety 2013. Geneva: WHO.

World Health Organization (WHO) (2014) Injuries and violence: The facts. Available at: https://apps.who.int/iris/bitstream/handle/10665/149798/9789241508032_eng.pdf

World Health Organization (WHO) (2020) Global health estimates: Leading causes of death. Available at: https://www.who.int/data/gho/data/themes/mortality-and-global-health-estimates/ghe-leading-causes-of-death

